# Comparative Analysis of Real–World Acute Prescription Migraine Therapy Outcomes: Insights from the HeAD–US Study

**DOI:** 10.1101/2025.09.18.25336093

**Authors:** Devin Teichrow, Babak Khorsand, Kristina M. Fanning, Alexandre Urani, François Cadiou, Richard B. Lipton, Ali Ezzati

**Affiliations:** Department of Neurology, University of California, Irvine (UCI), Irvine, USA; Mist Research, Wilmington, NC, USA; APTAR LLC, USA; Department of Neurology, Albert Einstein College of Medicine and Montefiore Medical center, Bronx, NY, USA

**Keywords:** Migraine, Treatment Optimization, Gepants, Triptans, Pain Relief, Treatment Patterns, Treatment Response

## Abstract

**Background:** Triptans have long served as the primary acute migraine treatment, while gepants represent a newer, non-vasoconstrictive alternative. Although clinical trials have explored efficacy within each class, head-to-head studies are lacking, and real-world data on optimal treatment response and patient-level predictors of outcomes remain limited. We leverage baseline data from the HeAD–US study to conduct real-world comparisons of triptans and gepants for 2-hour pain freedom (2hPF) and 24-hour pain relief (24hPR), and to identify demographic, clinical, and treatment-related predictors of these outcomes.

**Methods:** Head-US is a survey conducted among users of the Migraine Buddy application. Eligible participants completed the survey, met ICHD–3 criteria for migraine and reported using acute monotherapy with a gepants (n = 570) or triptan (n = 1018). Primary outcomes, 2hPF and 24hPR post-treatment, were assessed using the Migraine Treatment Optimization Questionnaire (mTOQ-6). Potential predictors included demographic factors, clinical measures, and treatment-related factors. Logistic regression models were applied to complete-case data, with pairwise comparisons estimated via marginal-means.

**Results:** Among 1,588 eligible respondents 570 used gepants and 1,018 used a triptan. Mean age was 43.35 years (SD = 13.04), and 94.8% were female. Gepant users demonstrated 39% higher odds of achieving 24hPR compared to triptan users (OR = 1.39, 95% CI: 1.11–1.74) though there were no significant differences the the 2hPF outcome. Reduced odds of adequate 2hPF were associated with higher migraine symptom severity, high-frequency episodic migraine, chronic migraine, severe pain intensity, and severe migraine disability. Odds of achieving 24hPR were reduced in those with higher migraine symptom severity, higher attack frequency, severe migraine disability, and preventive medication use. Predictors of treatment response were similar in the pooled sample and in those using gepants and in those using triptans.

**Conclusion:** In this large, real-world cohort, triptans and gepants showed comparable effectiveness for 2hPF, but gepants demonstrated a significant advantage for 24hPR. Clinical features such as symptom severity, headache frequency, disability, and comorbid treatment burden were important predictors of treatment response. These findings support the need for larger, head-to-head clinical trials definitively comparing these medication classes for migraine management and may inform personalized selection of acute migraine therapies in clinical practice.

## Introduction

Migraine, a highly prevalent and disabling neurological disorder, impacts one billion individuals worldwide^1^. It ranks as the second most disabling condition after stroke, contributing approximately 45.1 million disability–adjusted life years globally^2^. Despite the availability of multiple acute treatment options, many individuals continue to experience suboptimal symptom relief, delayed onset of action, or unacceptable side effects, highlighting the ongoing need for more effective and better–tolerated therapies^3^.

While over-the-counter (OTC) medications are the most widely used treatment for migraine, for decades, triptans (serotonin 5–HT□b/d receptor agonists) have served as the most widely used prescription pharmacologic treatment for acute migraine^4^. While generally effective, triptans can induce vasoconstriction, leading to contraindications for those with cardiovascular disease and precautions for those at risk for cardiovascular disease ^5^. The advent gepants, has expanded the therapeutic landscape as they offer a non–vasoconstrictive mechanism of action and have a better adverse event profile than triptans ^6,7^.

Recent clinical studies suggest that while triptans remain a first-line acute treatment for migraine, the newer gepant class offers a valuable alternative, especially for patients with cardiovascular contraindications or those who experience insufficient response or tolerability issues with triptans^8^. The comparative efficacy between these two classes for acute migraine remains an area of ongoing discussion. Some network meta-analyses have concluded that certain triptans demonstrate superior efficacy for 2–hour pain freedom (2hPF) and relief compared to gepants^6,9^. However, these analyses often pool trials sometimes conducted decades apart, with shifts in primary endpoints (e.g., from pain relief to pain freedom and most bothersome symptom relief) and changes in data capture methods (e.g., paper to electronic diaries), potentially influencing reported outcomes^8^. Furthermore, contemporary randomized trials have shown lower 2hPF rates for triptans than historically reported, raising questions about the generalizability of older efficacy data^10^. Conversely, real-world studies and some clinical observations suggest that gepants can achieve 2hPF rates comparable to triptans, with potential advantages in maintaining relief over longer periods, possibly due to their pharmacokinetics, and a favorable tolerability profile^11^. These complexities underscore the importance of real-world evidence to better understand the comparative effectiveness and guide personalized treatment strategies.

While direct comparisons of acute migraine treatments are crucial, an equally critical question is *which* patients are more or less likely to benefit from specific therapies. Previous studies have identified several demographic, clinical, and behavioral characteristics such as male sex, higher headache frequency, greater migraine–related disability, ictal–cutaneous allodynia, and depression as predictors of suboptimal response to acute treatments^12–16^. However, much of this prior work has focused on short–term (2–hour) outcomes or primarily examined triptans or non– specific analgesics, such as acetaminophen and ibuprofen. As a result, we have limited insight into the sustained efficacy of newer migraine–specific treatments across diverse patient populations, underscoring a significant gap in current literature.

To address these gaps, we leveraged data from the Headache Assessment via Digital Platform in the United States (HeAD–US), a large, app–based observational cohort of individuals with migraine. This study had two primary objectives: First, we examined real-world treatment outcomes for individuals using triptans or gepants as monotherapy, focusing on two commonly used endpoints. Second, we examined demographic, clinical, and treatment–related predictors of response to triptans and gepants specifically, with the goal of informing individualized treatment selection between these two classes. Based on the pharmacokinetic differences, particularly the extended half-lives of gepants compared to most used triptans (e.g. sumatriptan), we hypothesize that gepants will offer more sustained receptor occupancy. This leads us to predict that gepants will provide superior 24-hour pain relief (24hPR), with comparable efficacy to triptans at the 2-hour mark. We also hypothesized that patient–specific characteristics, such as greater headache burden, disability, and comorbidities, would predict reduced treatment response.

## Methods

### Study Design and Participants

This cross-sectional analysis used baseline data from the Headache Assessment via Digital Platform in the United States (HeAD–US), an observational study of adults with migraine who completed an app-based survey through Migraine Buddy. Details about the HeAD-US study are reported elsewhere^17^. In brief, over a three-month period beginning in September 2023, users of the app were invited to participate via in-app notification. Individuals consented electronically and completed a structured questionnaire assessing migraine history, treatment patterns, and related clinical characteristics. The survey gathered sociodemographic information, headache features such as severity and frequency, neuropsychiatric symptoms, use of acute and preventive treatments, and treatment responses. Quality control process included a quality control question, (also known as attention checks), which was used in the middle of a survey, and through verifying programmed response ranges and performing consistency checks. Response options like “prefer not to answer,” “don’t know,” “does not apply to me,” and “don’t remember” were included to accommodate participants who couldn’t or didn’t want to provide a definitive answer to specific questions, aiming to minimize missing data. Survey questions were presented in the same order to all participants, prioritizing essential questions. Adaptive question logic was applied where appropriate.

For this study, the sample was restricted to participants meeting ICHD–3 diagnostic criteria for migraine. Participants using opioid, barbiturate, and ergot–based treatments were excluded due to a small sample size, as well as those on over–the–counter (OTC) medications, due to confounding by indication if left in.

All participants provided electronic consent for participation in a general, survey–based migraine study and subsequently received the study questionnaire through the app. This study is a secondary analysis of deidentified data from the HeAD study, which was reviewed and approved by the Advarra, Inc. Institutional Review Board (CIRBI Protocol #Pro00072897). The secondary analysis described in the manuscript was reviewed and approved by the Institutional Review Board of the University of California, Irvine (IRB # 5459)

### Study Measures

The following demographics and clinical features were collected from all participants:

1. Demographics: Age, sex, and marital status
2. Headache features and symptoms: Includes headache pain intensity, Migraine Symptom Severity Score (MSSS) ^18^, average monthly headache days, and cutaneous allodynia. We included the validated AMS/AMPP Diagnostic Module to assess the proportion of participants who used the app who met criteria for migraine ^18^.
3. Depression and disability: Assessed using Depression and Anxiety (PHQ–4)^19^ and Migraine Disability Assessment (MIDAS) ^20^. A score of at least 4 was deemed indicative of at least mild symptoms of depression and anxiety. MIDAS scores were graded as none (scores: 0-5) to mild (6-10), moderate (11-20), and severe (21+),
4. Treatment patterns: Respondents reported their lifetime and current use of acute and preventive medications for migraine available at the time of the survey. Participants were asked separately about the classes of medications they use for acute or chronic treatment of migraine. Medication categories were provided, each listing several names of most common medications in each category, to capture the pharmacologic classes comprehensively.

### Primary Outcomes

The Migraine Treatment Optimization Questionnaire-6 (mTOQ-6) evaluates the effectiveness and optimization of migraine treatments through six questions. This tool helps healthcare providers assess patients’ responses to medication and identify areas for improvement. Like other recall-based questionnaires, mTOQ-6 is subject to subjective self-report and recall bias; however its items demonstrate high test-retest reliability and validity in clinical settings^21^. Treatment outcomes were defined using two individual items from the mTOQ-6: one assessing self-reported pain freedom within 2 hours of taking medication (2hPF), and another assessing whether pain relief was sustained at 24 hours post-treatment (24hPR). For the purposes of this study, treatment effectiveness was defined as an mTOQ-6 score of 6 or higher, corresponding to “moderate” (scores 6–7) or “maximum” (score 8) treatment effectiveness.

### Migraine Diagnosis

The “ICHD–based case definition for probable or definite migraine” was derived from participant responses to the AMS/AMPP diagnostic module^18^. This structured questionnaire was originally developed to align with ICHD–2 criteria and has since been validated for continued use under ICHD–3. Prior validation studies have shown that the module performs well against clinician–determined diagnoses, with a sensitivity of 100% and specificity of 82% for identifying migraine^18^.

### Statistical Analysis

All statistical analyses were conducted using RStudio (v. 4.3.1). Baseline demographic and clinical characteristics of the study cohort were summarized using descriptive statistics. Continuous variables were presented as means with standard deviations (SD), while categorical variables were reported as counts and percentages. These characteristics were further stratified by acute treatment group (triptans vs. gepants) to provide a comprehensive profile of the study population.

The primary outcomes (2hPF and 24hPR) were modeled as binary variables. Multivariable logistic regression was used to estimate associations between treatment response and medication class, adjusting for age, sex, MSSS, average monthly headache days (categorized as low-frequency episodic, high-frequency episodic, or chronic migraine), cutaneous allodynia, PHQ-4 score, MIDAS score, pain intensity (mild, moderate, severe), and preventive medication use. Fully adjusted models were used for both outcomes. To evaluate medication-specific effects, additional logistic regression models were stratified by medication class (triptans or gepants). For all models, coefficients were estimated on the log-odds scale and exponentiated to yield odds ratios (ORs) with 95% confidence intervals (CIs).

Model-based predicted probabilities were calculated using marginal standardization via the emmeans package in R. These marginal means were used to estimate absolute risk differences and risk ratios, with 95% confidence intervals derived using the delta method. All analyses were performed using RStudio (v4.3.1). The glm and emmeans packages were used for modeling and marginal effects estimation. Additional packages included margins and broom for model output formatting and visualization. Only participants with complete data for all covariates and outcomes were included in regression models (complete-case analysis); no imputation was performed.

## Results

### Sample Characteristics

Of the 6810 initial respondents to the questionnaire, 6606 (97.0%) met the ICHD–3 based case definition for probable or definite migraine. Of these, 5018 were removed due to being on multiple medications or OTC medications, leaving 1588 (24.0%) who met the ICHD–3 migraine case definition and were using either triptans (n = 1018) or gepants (n = 570). The flow of participant selection is summarized in Figure 1.

**Fig. 1.**
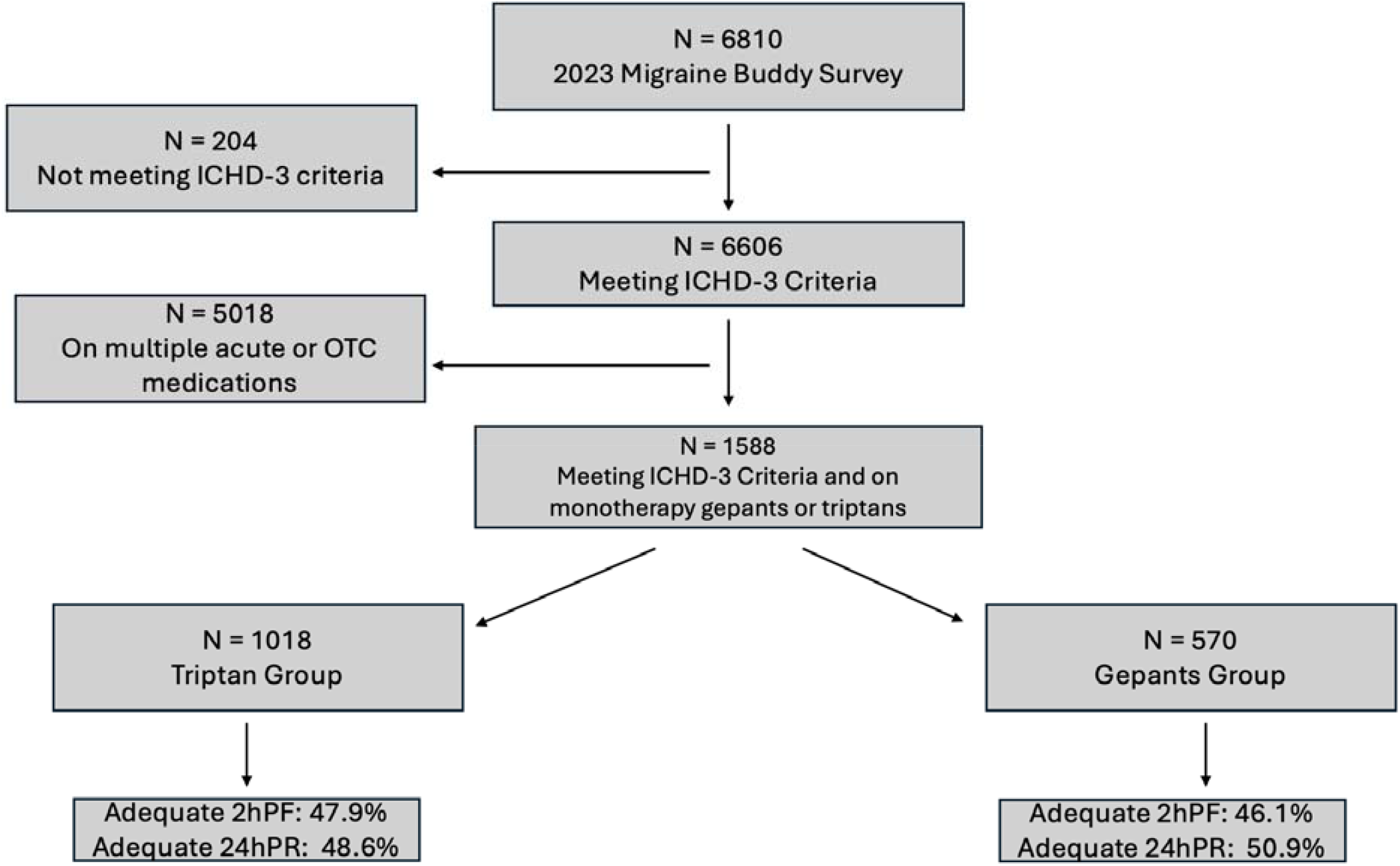
ICHD-3 = International Classification of Headache Disorders -3; OTC = over-the-counter; 2hPF = 2-hour pain freedom; 24hPR = 24-hour pain relief. Adequate response was defined using two individual items from the mTOQ-6: one assessing self-reported pain freedom within 2 hours of taking medication (2hPF): “After taking your migraine medication are you pain-free within 2 hours for most attacks?”, and another assessing whether pain relief was sustained at 24 hours post-treatment (24hPR): “Does one dose of your migraine medication usually releave your headache and keep it away for at least 24 hours?”. Responses of “half the time or more” were classified as successful outcomes for each respective domain.

Table 1 summarizes the demographic and clinical characteristics of the sample broken down by medication group. The cohort had an average age of 43.35 years (SD = 13.0), with 94.8% of participants being female. Chronic migraine was more prevalent in gepant users (38.4%) than in triptan users (27.7%). Moderate or severe pain intensity was common in both groups, and over 75% of participants met criteria for severe migraine-related disability. Preventive medication use was higher among gepant users (81.9%) than triptan users (71.2%).

**Table 1.**
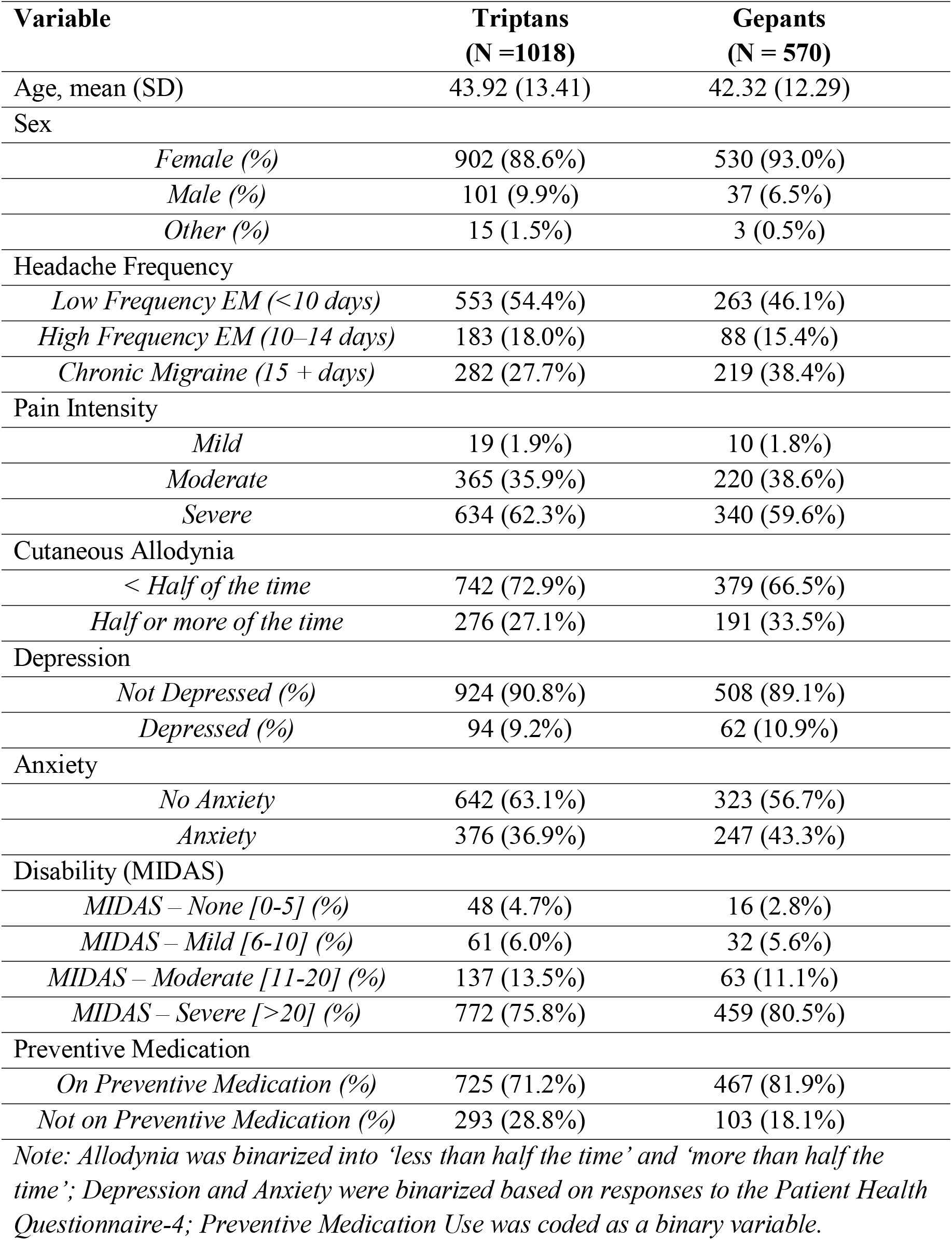
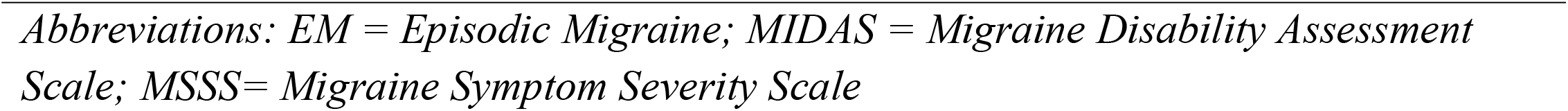
Demographics and Clinical Characteristics by Medication Class

**Table 2.** Overall Success* Rates for 2–Hour and 24–Hour Outcomes by Medication Group

| **Medication Class** | **Total** | **2hr Success**<br>**(n)** | **2hr Success**<br>**(%)** | **24hr Success**<br>**(n)** | **24hr Success (%)** |
| --- | --- | --- | --- | --- | --- |
| Triptan | 1018 | 488 | 47.9 | 495 | 48.6 |
| Gepants | 570 | 263 | 46.1 | 290 | 50.9 |

*\*Treatment outcomes were defined using two individual items from the mTOQ-6: one assessing self-reported pain freedom within 2 hours of taking medication (2hPF), and another assessing whether pain relief was sustained at 24 hours post-treatment (24hPR). Responses of “half the time or more” were classified as successful outcomes for each respective domain*

### Comparison of Treatment Response Between Gepants and Triptans

Overall, 47.9% participants using triptans experienced 2hPF and 48.6% achieved 24hPR. For the gepant group, 46.1% achieved a 2hPF while 50.9% achieved 24hPR.

In the fully adjusted logistic regression model for 2hPF (Table 3), there was no statistically significant difference between treatment classes (OR = 1.08, 95% CI: 0.86–1.34). Model-based predicted probabilities were 56.5% (95% CI: 47.6–65.0) for triptans and 58.3% (95% CI: 48.9– 67.1) for gepants, yielding an absolute difference of 1.8 percentage points and a non-significant risk ratio of 1.03.

**Table 3.** Multivariate Logistic Regression models for 2 hr pain freedom and 24hr pain relief in monotherapy users.

| Outcome | 2hr Pain Freedom |  |  | 24hr Sustained Pain Relief |  |  |
| --- | --- | --- | --- | --- | --- | --- |
| Variable | OR | LCL | UCL | OR | LCL | UCL |
| (Intercept) | 12.42 | 3.11 | 52.57 | 11.56 | 2.84 | 49.38 |
| Medication Class<br>(Reference = Triptans) |  |  |  |  |  |  |
| Gepants | 1.08 | 0.86 | 1.34 | **1.39** | **1.11** | **1.74** |
| Age | **1.01** | **1.01** | **1.02** | **1.01** | **1.00** | **1.02** |
| Sex (Reference =Female) | 1.09 | 0.81 | 1.45 | 1.04 | 0.77 | 1.39 |
| MSSS | **0.94** | **0.90** | **0.99** | **0.95** | **0.90** | **1.00** |
| Pain Frequency Per Month<br>(Reference =<br>< 10 days per month) |  |  |  |  |  |  |
| High Frequency EM (10–14 days) | **0.71** | **0.53** | **0.95** | **0.69** | **0.51** | **0.92** |
| Chronic Migraine ($\geq 15$ days) | **0.40** | **0.31** | **0.52** | **0.33** | **0.25** | **0.43** |
| Pain Intensity<br>(Reference = Mild Pain Intensity) |  |  |  |  |  |  |
| Moderate Pain Intensity | 0.51 | 0.20 | 1.20 | 0.78 | 0.31 | 1.82 |
| Severe Pain Intensity | **0.39** | **0.15** | **0.92** | 0.57 | 0.22 | 1.32 |
| Migraine Disability Assessment<br>Scale (MIDAS) –<br>(Reference = No burden and mild) |  |  |  |  |  |  |
| MIDAS – Moderate | **0.57** | **0.36** | **0.90** | **0.46** | **0.28** | **0.75** |
| MIDAS – Severe | **0.46** | **0.31** | **0.67** | **0.39** | **0.25** | **0.58** |
| Depression<br>(Reference = no depression) | 0.93 | 0.71 | 1.23 | 0.85 | 0.64 | 1.12 |
| Anxiety<br>(Reference = no anxiety) | 0.91 | 0.70 | 1.17 | 0.87 | 0.67 | 1.13 |
| Allodynia | 0.97 | 0.88 | 1.07 | 0.94 | 0.85 | 1.04 |
| Preventive Medication Use | 0.84 | 0.66 | 1.08 | **0.66** | **0.51** | **0.85** |

*Note: Allodynia, Depression, and Preventive Medication Use were coded as binary variables. Abbreviations: EM = Episodic Migraine; MIDAS = Migraine Disability Assessment Scale; MSSS= Migraine Symptom Severity Scale*

In contrast, gepant use was significantly associated with higher odds of achieving 24hPR (OR = 1.39, 95% CI: 1.11–1.74; Table 3). Marginal predicted probabilities were 54.5% (95% CI: 45.6– 63.1) for triptans versus 62.5% (95% CI: 53.2–70.9) for gepants. This corresponds to an absolute difference of 8.0 percentage points and a risk ratio of 1.15 (95% CI: 1.02–1.29).

### Predictors of Treatment Response

In the full logistic regression model for 2hPF (Table 3), several clinical factors emerged as statistically and clinically significant predictors of reduced response. In comparison to the lower frequency group, the odds of achieving 2hPF were significantly reduce for the high-frequency episodic migraine (HFEM) (OR = 0.71, 95% CI: 0.53–0.95), and the chronic migraine group (OR = 0.40, 95% CI: 0.31–0.52). Higher pain intensity was also a significant predictor of reduced response; severe pain intensity was associated with an OR of 0.39 (95% CI: 0.15–0.92, both compared to mild disability. Participants with severe disability on the MIDAS scale had significantly lower odds of achieving 2hPF compared to those with no or mild burden (OR = 0.46, 95% CI: 0.31–0.67). Other covariates, including age, sex, MSSS, allodynia, and depression severity, did not demonstrate either statistically or clinically meaningful associations with 2hPF after adjustment.

For the 24hPR outcome (Table 3), HFEM (OR = 0.69, 95% CI: 0.51–0.92) and chronic migraine (OR = 0.33, 95% CI: 0.25–0.43) were again associated with lower odds of success. Pain intensity was not significantly associated with 24hPR in the fully adjusted model. However, both moderate and severe disability significantly diminished the likelihood of achieving 24hPR. The OR for moderate disability was 0.46 (95% CI: 0.28–0.75), while for severe disability it was 0.39 (95% CI: 0.25–0.58). Preventive medication use was also associated with lower odds of 24hPR (OR = 0.66, 95% CI: 0.51–0.85).

Stratified logistic regression models by medication class are presented in Table 4. For 2hPF, higher migraine symptom severity was significantly associated with reduced response in both treatment groups. Among triptan users, the OR was 0.92 (95% CI: 0.86–0.98), and among gepant users, the OR was 0.93 (95% CI: 0.87–0.99). High-frequency episodic migraine and chronic migraine were associated with poorer 2hPF responses across both medication classes. In the triptan group, the ORs were 0.69 (95% CI: 0.48–0.99) for HFEM and 0.47 (95% CI: 0.34–0.65) for chronic migraine. In the gepants group, the corresponding ORs were 0.66 (95% CI: 0.46– 0.95) and 0.38 (95% CI: 0.27–0.53), respectively. Similarly, moderate and severe migraine-related disability were associated with reduced odds of 2hPF in both groups.

**Table 4.**
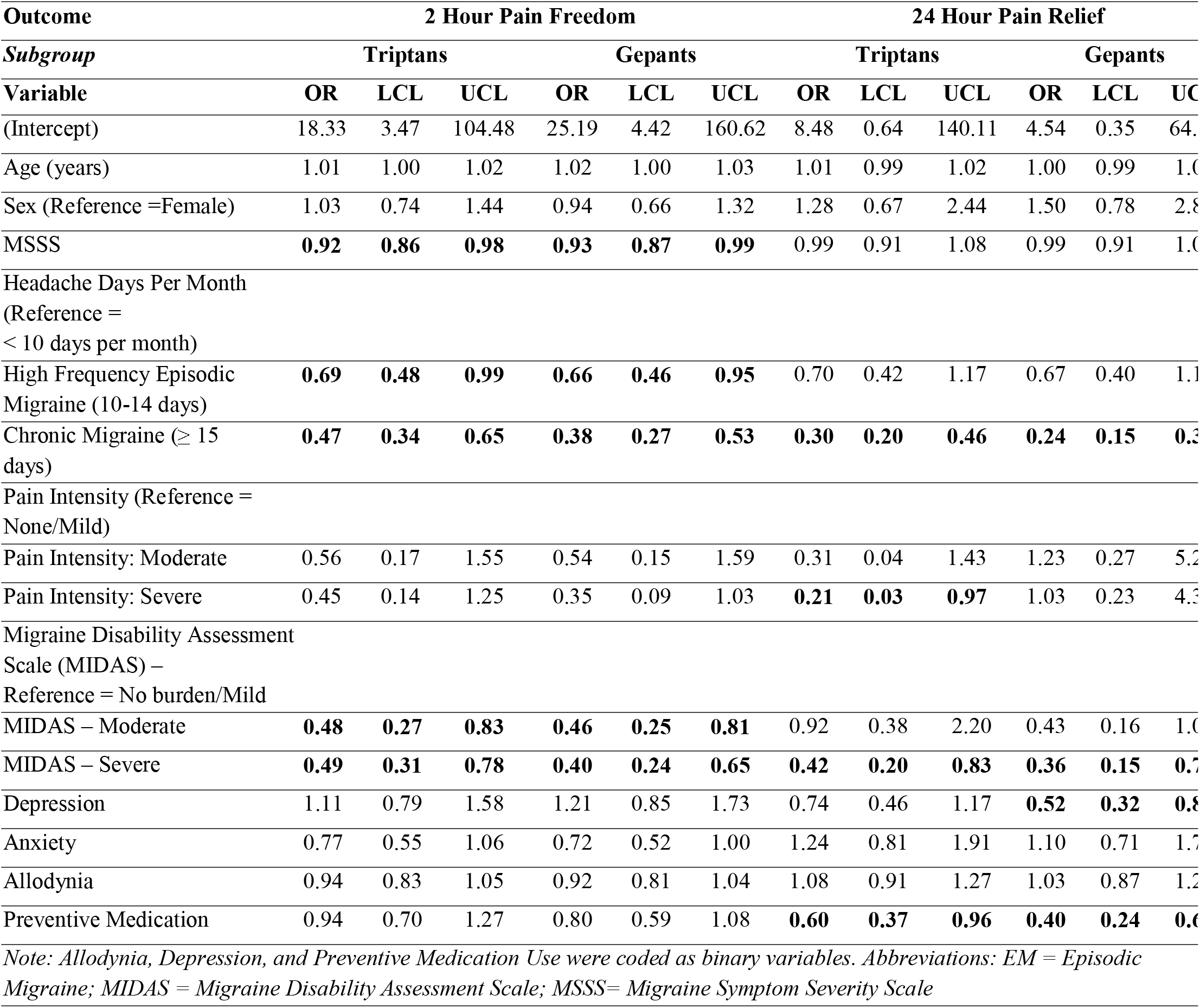
Logistic Regression models for 2hPF and 24hPR stratified by medication class

For 24hPR, chronic migraine was the only headache frequency variable that consistently predicted reduced response in both treatment groups. In the triptan group, the OR for chronic migraine was 0.30 (95% CI: 0.20–0.46), while in the gepant group it was 0.24 (95% CI: 0.15– 0.37). Severe migraine-related disability was associated with lower odds of 24hPR in both triptan users (OR = 0.42, 95% CI: 0.20–0.83) and gepant users (OR = 0.36, 95% CI: 0.15–0.77). Depression was associated with reduced 24hPR among gepant users (OR = 0.52, 95% CI: 0.32– 0.84), but not among triptan users. Severe pain intensity was significantly associated with lower odds of 24hPR in the triptan group (OR = 0.21, 95% CI: 0.03–0.97), but not in the gepant group. Preventive medication use was associated with lower 24hPR in both groups, with an OR of 0.60 (95% CI: 0.37–0.96) in the triptan group and 0.40 (95% CI: 0.24–0.66) in the gepant group.

## Discussion

In this large, real-world cohort of adults using acute monotherapy for migraine, we found no significant difference between gepants and triptans in 2hPF, while gepants were associated with higher odds of pain relief at 24 hours. We also identified several patient-level characteristics, including migraine frequency, pain intensity, and disability, that predicted poor response across outcomes.

While prior clinical trials have demonstrated the efficacy of both gepants and triptans for acute migraine treatment, these trials were generally conducted in different time periods, with differing eligibility criteria and endpoints. Recent network meta-analyses, such as that by Karlsson et al.^8^, have ranked triptans higher than gepants in relative efficacy. However, those rankings are based on indirect comparisons across separate trials, and are subject to potential confounding by trial design, population differences, and decade of conduct^9^. The triptan trials included in the Karlsson review were mostly conducted in the early 2000s, whereas gepant trials were conducted more recently. Thus, while useful for broad comparisons, these analyses are not substitutes for head-to-head comparative data. This observational study serves to provide some evidence while we await the results of randomized trials. In our study, using a real-world cohort and standardized outcome definitions, we found no significant difference in 2-hour pain freedom, but gepants were associated with a higher probability of achieving 24-hour relief, even after adjustment for multiple confounders.

This advantage may reflect known pharmacokinetic differences between the drug classes. Gepants, such as rimegepant (11 hour half-life) and ubrogepant (5-6 hour half life) have longer half lives that the most widely used triptans, sumatriptan and rizatriptan (1.7–2.2 hours), with the longer half-life triptans are less widely ^7, 10^. The longer half-life may result in more durable symptom relief, supporting their potential role for patients with longer headaches who prioritize sustained efficacy in acute migraine treatment.

We also identified several predictors of reduced response across both 2-hour and 24-hour outcomes. We found that higher migraine frequency, greater pain intensity, and severe migraine-related disability were consistently associated with reduced likelihood of treatment success at both 2 and 24 hours. These results are consistent with prior literature indicating that individuals with more severe or burdensome migraine profiles are less likely to respond to acute medications^11-13^. Preventive medication use was also associated with lower odds of 24-hour response, likely reflecting confounding by indication due to a higher baseline disease burden.^22,23^ Depression predicted reduced 24hPR among gepant users, but not triptan users, raising questions about potential interaction between treatment class and comorbidity that merit further study.

These findings have practical implications for treatment selection in clinical practice. While triptans remain a widely used and effective first-line option, gepants offer a non-vasoconstrictive alternative that may be preferable for individuals with cardiovascular contraindications, poor triptan tolerability, or a need for longer-lasting relief. Although gepants were associated with significantly greater odds of achieving 24-hour pain relief compared with triptans, the absolute difference in predicted probabilities was modest (about 8 percentage points). This suggests that both medication classes remain broadly effective, with gepants offering a small but clinically meaningful advantage in sustained relief. However, gepants are generally newer, more costly, and less widely accessible than triptans, and insurance coverage may vary. Thus, treatment decisions should balance clinical characteristics, patient preferences, safety considerations, and access to therapies to optimize individualized care.

Our study has several strengths, including its large sample size, real-world setting, and use validated outcome measures. By focusing on monotherapy users, we were able to isolate effects by medication class without the confounding of combination regimens. However, several limitations should be noted. First, the use of self–reported data on medication usage and treatment outcomes introduces the possibility of recall bias or misclassification bias, as we did not have objective verification of reported information. Although medication use and treatment outcomes were self-reported, pain relief is inherently subjective and patient-reported outcome measures (e.g., mTOQ-6) are standard in migraine research. If recall error is largely nondifferential by drug class, it would typically bias estimates toward the null. In addition, treatment groups were determined by the clinical prescribing process. This observational study does not have the benefit of random allocation of patients to treatment. Generally, patients who receive gepants have failed one or more triptans or ineligible for triptans. This form of paitient selection might be expected to favor tripans over gepants, the opposite of the results we found. Notably, we still observed significant differences between gepants and triptans, which supports the credibility of our approach. Second, the recruitment method may underrepresent certain demographics, such as less technologically informed older adults, individuals with reduced digital access, and those from lower socioeconomic backgrounds. Therefore, findings may not be fully generalizable to the broader population of individuals with migraine. Third, due to recruitment having been conducted in the app via a pop–up message, we are unable to determine how many users saw the invitation or clicked on it. We acknowledge this is a possible source of selection bias, as those who clicked on the pop–up may have differed from those who did not engage. However, this possibility exists in all survey–based research, as only a small proportion of individuals who are contacted typically follow through to participation. In addition, because this study was cross-sectional, it cannot capture dose- or timing-related effects, long-term treatment effectiveness, adherence, or changes in treatment patterns over time. Lastly, recruitment through a smartphone app may preferentially engage younger, more digitally connected individuals, which could limit the representativeness of our findings for the broader migraine population. However, the sample’s mean age (43 years) is comparable to prior trials and surveys in migraine, which mitigates concerns about age-specific selection. Generalizability along other dimensions (e.g., socioeconomic status, access to care) may still be limited.

In conclusion, this real-world analysis found that gepants and triptans performed similarly for 2hPF, while gepants offered a modest advantage for 24hPR. Clinical features such as headache frequency, disability, and pain severity were associated with lower response across both treatment classes. These findings contribute to a more nuanced understanding of acute migraine treatment effectiveness and support more personalized approaches to care.

## Data Availability

Data available upon reasonable request.

## List of Abbreviations

2hPF: 2–hour pain freedom
24hPR: 24–hour pain relief
AMS/AMPP: American Migraine Study/ American Migraine Prevalence and Prevention
CI: Confidence Interval
EMM: Estimated Marginal Means
HeAD–US: Headache Assessment Via Digital Platform in United States
ICHD: International Classification of Headache Disorders
MIDAS: Migraine Disability Assessment
MSSS: Migraine Symptom Severity Scale
mTOQ: Migraine Treatment Optimization Questionnaire
NMA: Network Meta–analysis
OR: Odds Ratio
PHQ–4: Patient Health Questionnaire for Depression and Anxiety
SD: *Standard Deviation*

## Ethics Approval and Consent to Participate

Institutional Review Board (CIRBI Protocol # Pro 00072897)

## Availability of Data and Materials

The datasets used and analyzed during the current study are available from the corresponding author upon reasonable request.

## Competing Interests

Ali Ezzati receives research support from the following sources: National Institute of Health (NIA K23 AG063993; NIA– 1R01AG080635–01A1); the Alzheimer’s Association (SG–24– 988292), Cure Alzheimer’s Fund, and Amgen investigator–initiated studies.

Kristina M Fanning is the managing director of MIST Research, LLC which received grants from the National Headache Foundation in addition to funding from Allergan, Amgen, Dr. Reddy’s Laboratories/Promius, and Eli Lilly via collaboration with Vedanta Research.

Alexandre Urani and François Cadiou are employees of APTAR LLC, which is the parent company of Migraine Buddy.

Richard B. Lipton receives research support from the NIH and the FDA as well as the National Headache Foundation and the Marx Foundation. He serves on the editorial board of Neurology, senior advisor to Headache, and associate editor to Cephalalgia. He has reviewed for the NIA and NINDS, holds stock options in Axon, Biohaven Holdings, CoolTech and Manistee; serves as consultant, advisory board member, or has received honoraria from: Abbvie (Allergan), American Academy of Neurology, American Headache Society, Amgen, Avanir, Biohaven, Biovision, Boston Scientific, CoolTech, Dr. Reddy’s (Promius), Electrocore, Eli Lilly, eNeura Therapeutics, GlaxoSmithKline, Grifols, Lundbeck (Alder), Pfizer, Teva, Trigemina, Vector, Vedanta. He receives royalties from Wolff’s Headache 7^th^ and 8^th^ Edition, Oxford Press University, 2009, Wiley and Informa.

## Funding

No funding was received for this Research.

## Author’s Contributions

**Devin Teichrow:** Conceptualization; investigation; methodology; formal analysis; writing – original draft; writing - review and editing

**Babak Khorsand:** Methodology, writing – original draft; writing – review and editing.

**Kristina M. Fanning:** Conceptualization; data curation; formal analysis; investigation; methodology; resources; writing – review and editing.

**François Cadiou:** Data curation; methodology; resources; software; writing – review and editing.

**Alexandre Urani**: Data curation; methodology; resources; software; writing – review and editing.

**Richard B. Lipton**: Conceptualization; data curation; investigation; methodology; project administration; resources; supervision; writing – review and editing.

**Ali Ezzati**: Conceptualization; data curation; investigation; methodology; resources; supervision; validation; visualization; writing – review and editing.

## Notes

### Funding Statement

This study did not receive any funding.

### Author Declarations

reviewed and approved by the Advarra, Inc. Institutional Review Board (CIRBI Protocol #Pro00072897). The secondary analysis described in the manuscript was reviewed and approved by the Institutional Review Board of the University of California, Irvine (IRB # 5459)

## References

1. Zhang N, Robbins MS. Migraine. Ann Intern Med. 2023;176(1):ITC1–ITC16. doi:10.7326/AITC202301170

2. Collaborators G 2019 A. Global, regional, and national burden of diseases and injuries for adults 70 years and older: systematic analysis for the Global Burden of Disease 2019 Study. BMJ. 2022;376:e068208. doi:10.1136/bmj-2021-068208

3. Eigenbrodt AK, Ashina H, Khan S, et al. Diagnosis and management of migraine in ten steps. Nat Rev Neurol. 2021;17(8):501–514. doi:10.1038/s41582-021-00509-5

4. Johnston MM, Rapoport AM. Triptans for the Management of Migraine. Drugs. 2010;70(12):1505–1518. doi:10.2165/11537990-000000000-00000

5. Hall GC, Brown MM, Mo J, MacRae KD. Triptans in migraine. Neurology. 2004;62(4):563–568. doi:10.1212/01.WNL.0000110312.36809.7F

6. Yang CP, Liang CS, Chang CM, et al. Comparison of New Pharmacologic Agents With Triptans for Treatment of Migraine. JAMA Netw Open. 2021;4(10):e2128544. doi:10.1001/jamanetworkopen.2021.28544

7. Negro A, Martelletti P. Gepants for the treatment of migraine. Expert Opin Investig Drugs. 2019;28(6):555–567. doi:10.1080/13543784.2019.1618830

8. Lipton RB, Gendolla A, Abraham L, et al. Relative frequency, characteristics, and disease burden of patients with migraine unsuitable for triptan treatment: A systematic literature review. Headache. 2025;65(1):164–179. doi:10.1111/head.14854

9. Karlsson WK, Ostinelli EG, Zhuang ZA, et al. Comparative effects of drug interventions for the acute management of migraine episodes in adults: systematic review and network meta-analysis. Published online September 18, 2024. doi:10.1136/bmj-2024-080107

10. Lipton RB, Goadsby PJ. Re: Comparative effects of drug interventions for the acute management of migraine episodes in adults: systematic review and network meta-analysis. Published online July 23, 2025. Accessed July 23, 2025. https://www.bmj.com/content/386/bmj-2024-080107/rr-4

11. Iannone LF, Vaghi G, Sebastianelli G, et al. Effectiveness and tolerability of rimegepant in the acute treatment of migraine: a real-world, prospective, multicentric study (GAINER study). J Headache Pain. 2025;26(1):4. doi:10.1186/s10194-024-01935-8

12. Ezzati A, Buse DC, Fanning KM, Reed ML, Martin VT, Lipton RB. Predictors of treatment-response to acute prescription medications in migraine: Results from the American Migraine Prevalence and Prevention (AMPP) Study. Clin Neurol Neurosurg. 2022;223:107511. doi:10.1016/j.clineuro.2022.107511

13. Ezzati A, Fanning KM, Buse DC, et al. Predictive models for determining treatment response to nonprescription acute medications in migraine: Results from the American Migraine Prevalence and Prevention Study. Headache J Head Face Pain. 2022;62(6):755–765. doi:10.1111/head.14312

14. Ezzati A, Fanning KM, Reed ML, Lipton RB. Predictors of treatment□response to caffeine combination products, acetaminophen, acetylsalicylic acid (aspirin), and nonsteroidal anti□inflammatory drugs in acute treatment of episodic migraine. Headache J Head Face Pain. 2023;63(3):342–352. doi:10.1111/head.14459

15. Lipton RB, Munjal S, Buse DC, Fanning KM, Bennett A, Reed ML. Predicting Inadequate Response to Acute Migraine Medication: Results From the American Migraine Prevalence and Prevention (AMPP) Study. Headache J Head Face Pain. 2016;56(10):1635–1648. doi:10.1111/head.12941

16. Porreca F, Navratilova E, Hirman J, van den Brink AM, Lipton RB, Dodick DW. Evaluation of outcomes of calcitonin gene-related peptide (CGRP)-targeting therapies for acute and preventive migraine treatment based on patient sex. Cephalalgia Int J Headache. 2024;44(3):3331024241238153. doi:10.1177/03331024241238153

17. Ezzati A, Fanning KM, Teichrow D, Urani A, Cadiou F, Lipton RB. Headache Assessment via Digital Platform in United States: Baseline study characteristics, diagnosis, and treatment patterns. Headache. Published online July 15, 2025. doi:10.1111/head.15016

18. Lipton RB, Diamond S, Reed M, Diamond ML, Stewart WF. Migraine Diagnosis and Treatment: Results From the American Migraine Study II. Headache J Head Face Pain. 2001;41(7):638–645. doi:10.1046/j.1526-4610.2001.041007638.x

19. Kroenke K, Spitzer RL, Williams JBW, Löwe B. An Ultra-Brief Screening Scale for Anxiety and Depression: The PHQ–4. Psychosomatics. 2009;50(6):613–621. doi:10.1016/S0033-3182(09)70864-3

20. Development and testing of the Migraine Disability Assessment (MIDAS) Questionnaire to assess headache-related disability. Neurology. Accessed March 3, 2025. https://www.neurology.org/doi/10.1212/WNL.56.suppl_1.S20

21. Lipton RB, Kolodner K, Bigal ME, et al. Validity and reliability of the Migraine-Treatment Optimization Questionnaire. Cephalalgia Int J Headache. 2009;29(7):751–759. doi:10.1111/j.1468-2982.2008.01786.x

22. Diamond S, Bigal ME, Silberstein S, Loder E, Reed M, Lipton RB. Patterns of Diagnosis and Acute and Preventive Treatment for Migraine in the United States: Results from the American Migraine Prevalence and Prevention Study: CME. Headache J Head Face Pain. 2007;47(3):355–363. doi:10.1111/j.1526-4610.2006.00631.x

23. Lipton RB, Munjal S, Alam A, et al. Migraine in America Symptoms and Treatment (MAST) Study: Baseline Study Methods, Treatment Patterns, and Gender Differences. Headache J Head Face Pain. 2018;58(9):1408–1426. doi:10.1111/head.13407

